# Early Impact of COVID-19 on Individuals with Eating Disorders: A survey of ~1000 Individuals in the United States and the Netherlands

**DOI:** 10.1101/2020.05.28.20116301

**Authors:** Jet D. Termorshuizen, Hunna J. Watson, Laura M. Thornton, Stina Borg, Rachael E. Flatt, Casey M. MacDermod, Lauren E. Harper, Eric F. van Furth, Christine M. Peat, Cynthia M. Bulik

## Abstract

We received rapid ethical permission to evaluate the early impact of COVID-19 on people with eating disorders. Participants in the United States (US, N=511) and the Netherlands (NL, N=510), recruited through ongoing studies and social media, completed an online baseline survey that included both quantitative measures and free-text responses assessing the impact of COVID-19 on situational circumstances, eating disorder symptoms, eating disorder treatment, and general well-being. Results revealed strong and wide-ranging effects on eating disorder concerns and illness behaviors that were consistent with diagnoses. Participants with anorexia nervosa (US 62% of sample; NL 69%) reported increased restriction and fears about being able to find foods consistent with their meal plan. Individuals with bulimia nervosa and binge-eating disorder (US 30% of sample; NL 15%) reported increases in their binge-eating episodes and urges to binge. Respondents noted marked increases in anxiety since 2019 and reported greater concerns about the impact of COVID-19 on their mental health than physical health. Although many participants acknowledged and appreciated the transition to telehealth, limitations of this treatment modality for this population were raised. Individuals with past histories of eating disorders noted concerns about relapse related to COVID-19 circumstances. Encouragingly, respondents also noted positive effects including greater connection with family, more time for self-care, and motivation to recover.

---

The novel coronavirus pandemic (COVID-19) has far-reaching effects on both physical and mental health. Many aspects of the pandemic are already adversely influencing mental health—the disease itself, restrictive policies aimed at reducing the spread of COVID-19, economic consequences, and more. A third of Americans were showing signs of clinical anxiety or depression by late May 2020.^1^ Addressing mental health needs is an integral part of the COVID-19 response.^2^ Factors related to COVID-19 will affect mental health in the general community,^3^ but may have greater adverse effects on individuals with pre-existing mental illnesses.^4-6^ Reports from China suggest that those with psychiatric illnesses before COVID-19 experienced a worsening in depression, anxiety, and post-traumatic stress symptoms during the peak of the outbreak.^7-10^

Individuals with eating disorders may face unique risks secondary to the pandemic. Public health measures designed to flatten the curve and the impact of COVID-19 on food availability can directly and adversely affect the core symptoms of eating disorders, including dietary restriction, binge eating, and compensatory behaviors. Narrative reviews and interviews with patients have highlighted issues such as intense urges to binge in those with binge-eating disorder (BED) when high risk foods are being stockpiled to guard against disruptions to the food supply. Individuals with some eating disorders such as anorexia nervosa (AN) or avoidant/restrictive food intake disorder (ARFID) may only be comfortable eating a very limited variety of foods, many of which may be brand-specific, but limited stock may prevent them from obtaining these foods altogether. Additionally, changes in the availability of treatment and social supports could hamper treatment progress or precipitate relapse, and comorbid psychiatric conditions (e.g., depression, anxiety) may worsen in the context of isolation and/or the economic impact of this pandemic.

The extent to which eating disorders may be affected by COVID-19 remains unknown. Specialist clinicians have highlighted the importance of attending to the unique needs of patients with eating disorders during this time^11-13^ and introduced potential adaptations to existing evidence-based interventions in inpatient^14^ and outpatient^15,16^ levels of care. However, the need for data-driven information from individuals with lived experience is urgent to guide clinicians in how best to deliver needed services.

One small study^11^ surveyed 32 patients with eating disorders in Spain about the impact of physical distancing measures during the first two weeks of confinement. They also reported qualitative data from a multifamily chat group in the United Kingdom consisting of eight patients with AN and their carers.^11^ Patients in both groups reported exacerbation of their eating disorder symptoms, increased anxiety, and challenges associated with reduced contact with their treatment teams, suggesting that patients with eating disorders may be at risk for symptom exacerbation and/or difficulty maintaining treatment progress during the global pandemic.

The objective of this study was to characterize the impact of COVID-19 on patients with eating disorders and to describe their treatment needs. Resultant data will inform best practices for clinicians and caregivers and provide a roadmap for eating disorders care as the pandemic evolves.

## METHODS

### Participants and procedure

Participants from the United States (US) and the Netherlands (NL) were invited to take part in an online study. US participants were recruited via social media (e.g., Facebook, Twitter, Instagram advertisements, and the UNC Exchanges blog), or via emails to participants who consented to recontact from ongoing studies.^17,18^ NL participants were also recruited via social media and via the online platform Proud2Bme and the Dutch Eating Disorder Register (NER). The minimum age to participate without parental consent was 18 years in the US and 16 years in NL. Participants were onboarded during a 29 day period from April 8^th^ up to and including May 6^th^ 2020 in the US, and from April 17^th^ up to and including May 15^th^ in NL. Consenting participants completed an online baseline survey and agreed to be recontacted approximately monthly on twelve occasions over the course of a year. The present study presents the results of the baseline survey. Ethical approval was granted by the University of North Carolina Biomedical Institutional Review Board. The Medical Research Ethics Committee (MREC-LDD) of Leiden University Medical Centre reviewed the study protocol and confirmed that the Medical Research Involving Human Subjects Act (WMO) did not apply to this study and official approval of this study by the METC was not required.

### Measures

The online survey addressed concerns and challenges participants are facing with regard to their eating disorder and general mental health during the COVID-19 global pandemic. The survey was developed in English, and translated to Dutch (see Supplementary Material).

#### Sociodemographics and illness status

Participants reported their age, sex, gender identity, and geographic location (US participants). Lifetime eating disorder diagnoses and current illness status were self-reported.

#### COVID-19 exposure and situational circumstances

Several questions captured COVID-19 exposure (exposure to the virus, diagnosis in self or family, and impact of COVID-19 on family members’ health and/or employment) and level of lockdown such as quarantine, physical distancing, voluntary or mandatory isolation, working from home, and shelter-in-place/stay-at-home orders.

#### Impact of COVID-19 on eating disorders

Questions about COVID-19-related concerns on eating disorders captured the previous two-week timeframe. A 4-point Likert-scale asked participants to rate their level of concern about access to and affordability of food and treatment, and worsening of eating disorder symptoms due to a lack of structure, changes in the social support environment, and time spent living in a triggering environment. The survey also assessed the impact of COVID-19 on specific eating disorder symptoms (e.g., “I have binged on food that I [or my family] have stockpiled”). These items were combined to create a mean impact score (Cronbach’s α = 0.77). Free-text items queried details on additional eating disorder-related concerns and any positive changes in their eating disorder or symptoms associated with the pandemic (if applicable).

#### Impact of COVID-19 on general physical and mental well-being

A 7-point Likert scale addressed the extent to which participants were worried about exposure to and/or contracting COVID-19. Relevant domains included: worry about self or others, worry about physical and/or mental health, and changes in overall anxiety levels since the start of the pandemic. Principal components analysis suggested these items loaded onto one component, so a total worry score was calculated (Cronbach’s α = 0.79). The Generalized Anxiety Disorder 7-Item Scale (GAD-7)^19^ assessed anxiety symptoms, and has demonstrated excellent psychometric properties.^20-23^ A score ≥ 10 is a positive screen for GAD.^19^ The survey also assessed whether participants had experienced a change in their levels of anxiety since the end of 2019, whether this change was due to COVID-19, and any positive changes in their lives related to COVID-19 circumstances.

#### Impact of COVID-19 on eating disorder treatment

We assessed COVID-19-related changes to eating disorder treatment modality, and for those currently receiving treatment, the frequency of sessions/contacts and perception of the quality of treatment. One free-text item inquired about the participants’ perceived greatest treatment needs.

### Statistical analysis

Data from the US and NL were analyzed separately to enable cross-country comparisons. We first report descriptives of the demographics, clinical characteristics, and COVID-19 exposure and situational circumstances. We assessed the different sections (effects of COVID-19 on eating disorder, eating disorder treatment, and general physical and mental wellbeing) by a descriptive analysis of the quantitative survey items. We then conducted one-way analyses of covariance (ANCOVA) to explore differences between groups (defined later) on continuous items from the various questionnaire sections, controlling for biological sex (male or female, intersex excluded), age, and day of study enrollment. Logistic regression was used for nominal variables. We used the following category definitions for group comparisons: (a) hierarchical lifetime diagnostic categories (i.e., a person can only be in one category) (AN, BN, BED, other eating disorders [OED]) in currently ill participants; (b) access to treatment (experiencing difficulties accessing one’s treatment provider versus not having difficulties); (c) eating disorder status (currently ill versus past eating disorder); and (d) level of lockdown. Free-text items were reviewed by three independent clinicians and summarized into global themes. As the *a priori* goals of the study were to conduct a broad analysis of areas of concern of patients with eating disorders in order to inform providers globally about healthcare service needs, we intentionally did not correct for multiple comparisons.

## RESULTS

### QUANTITATIVE RESULTS

#### Sociodemographic and illness characteristics

Table 1 presents demographic information for the US (*N* = 511) and NL (*N* = 510) samples, which were very similar. The mean age was young adult, ranging from 16 to over 60, and participants were predominantly female, and currently ill or symptomatic. US participants were well-distributed from across the country. In both countries, the majority of participants reported a diagnosis of AN. The next most common lifetime diagnoses were BN, BED, atypical AN, and other specified feeding or eating disorder/eating disorder not otherwise specified (OSFED/EDNOS), with different distributions between countries. Less common were ARFID, purging disorder, and night-eating syndrome. A full 56% of US and 34% of NL participants reported multiple lifetime eating disorder diagnoses, consistent with the known diagnostic fluctuation of these disorders.^24^ Slightly over half of participants in both samples reported being in treatment when COVID-19 struck. We excluded sex as a covariate for NL analyses, because of the small number (*N* = 4) male participants.

**Table 1.**
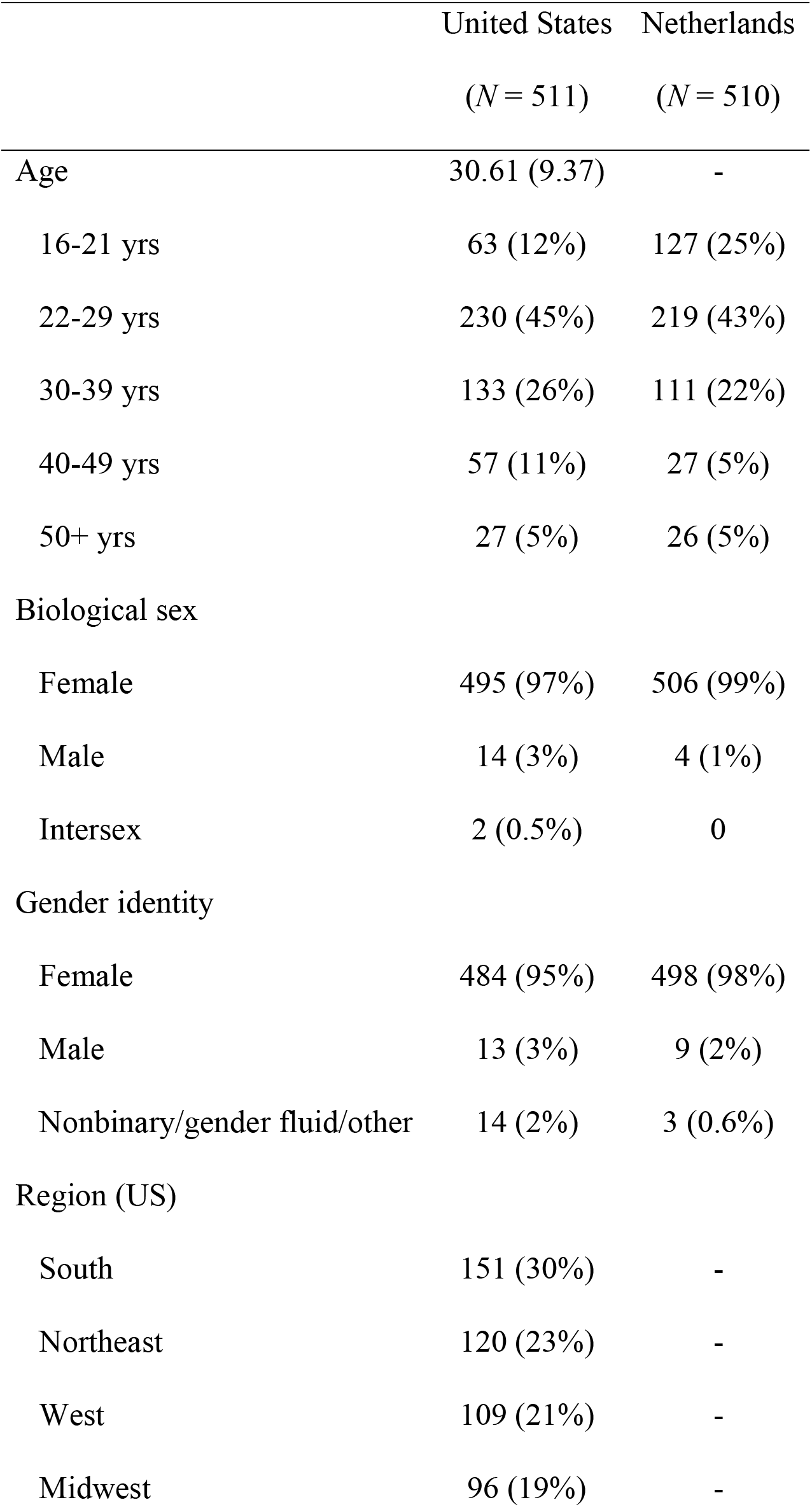
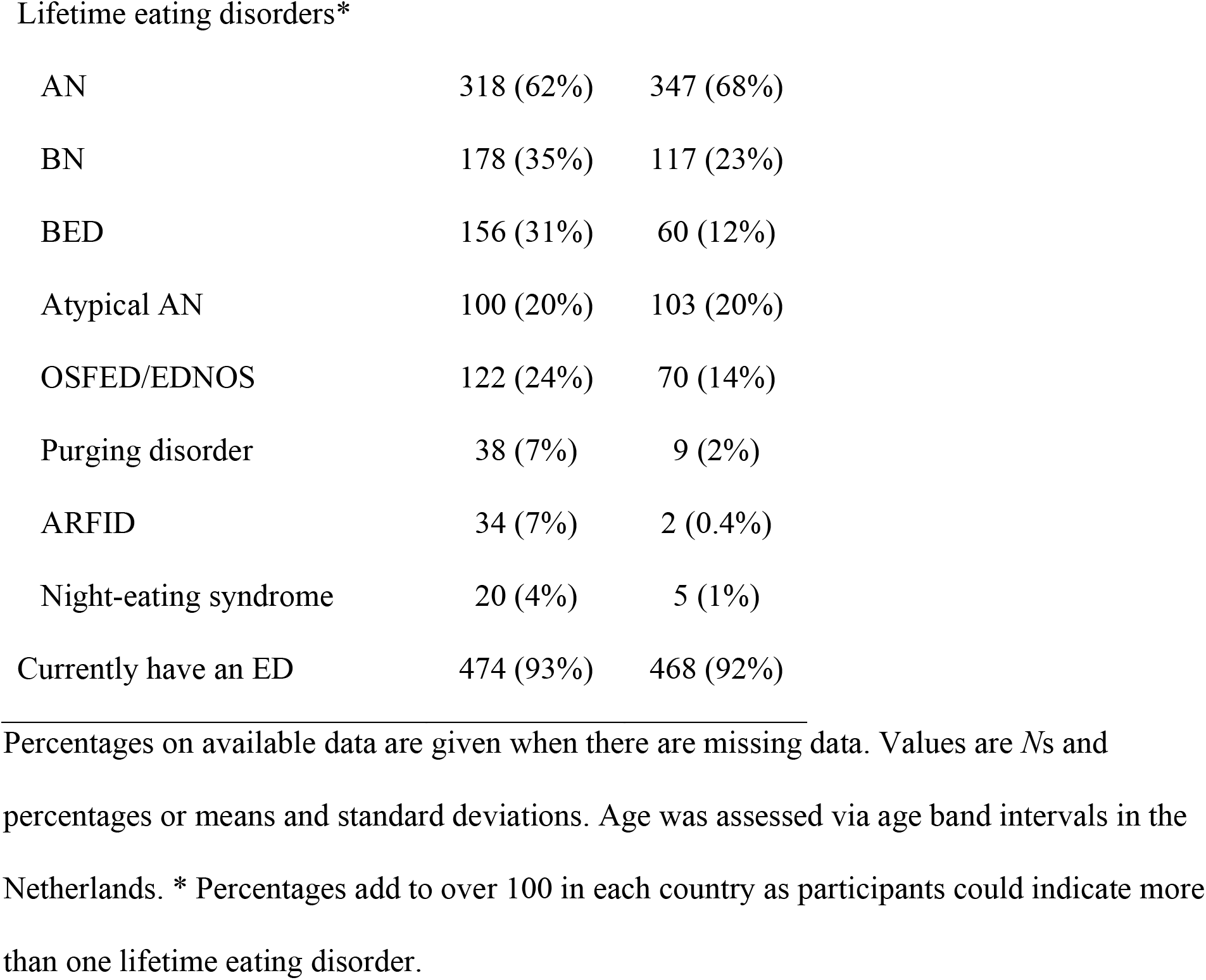
Sociodemographic and illness characteristics of the study population

|  | United States<br>(*N* = 511) | Netherlands<br>(*N* = 510) |
| --- | --- | --- |
| Age | 30.61 (9.37) | - |
| 16-21 yrs | 63 (12%) | 127 (25%) |
| 22-29 yrs | 230 (45%) | 219 (43%) |
| 30-39 yrs | 133 (26%) | 111 (22%) |
| 40-49 yrs | 57 (11%) | 27 (5%) |
| 50+ yrs | 27 (5%) | 26 (5%) |
| Biological sex |  |  |
| Female | 495 (97%) | 506 (99%) |
| Male | 14 (3%) | 4 (1%) |
| Intersex | 2 (0.5%) | 0 |
| Gender identity |  |  |
| Female | 484 (95%) | 498 (98%) |
| Male | 13 (3%) | 9 (2%) |
| Nonbinary/gender fluid/other | 14 (2%) | 3 (0.6%) |
| Region (US) |  |  |
| South | 151 (30%) | - |
| Northeast | 120 (23%) | - |
| West | 109 (21%) | - |
| Midwest | 96 (19%) | - |
| AN | 318 (62%) | 347 (68%) |
| BN | 178 (35%) | 117 (23%) |
| BED | 156 (31%) | 60 (12%) |
| Atypical AN | 100 (20%) | 103 (20%) |
| OSFED/EDNOS | 122 (24%) | 70 (14%) |
| Purging disorder | 38 (7%) | 9 (2%) |
| ARFID | 34 (7%) | 2 (0.4%) |
| Night-eating syndrome | 20 (4%) | 5 (1%) |
| Currently have an ED | 474 (93%) | 468 (92%) |

Percentages on available data are given when there are missing data. Values are *N*s and percentages or means and standard deviations. Age was assessed via age band intervals in the Netherlands. \* Percentages add to over 100 in each country as participants could indicate more than one lifetime eating disorder.

#### COVID-19 exposure

Table 2 presents information on extent of exposure to COVID-19. In the US, virtually all participants had been impacted by COVID-19 with 99% reported practicing physical distancing. Only 1% reported having a COVID-19 diagnosis, but many more reported some degree of self-isolation, quarantine, or working from home. Seven percent of respondents reported family members who had been infected and 25% reported a family member having lost employment due to COVID-19. The Dutch sample showed a similar pattern—98% reported practicing physical distancing and between one third and half of the participants reported voluntary self-isolation or working from home. In NL, 2% of the respondents reported having a COVID-19 diagnosis, 13% reported family members who had become physically ill, and 6% reported a family member having lost employment.

**Table 2.** COVID exposure of the study population

|  | United States | Netherlands |
| --- | --- | --- |
|  | (*N* = 511) | (*N* = 510) |
| COVID-19 diagnosis | 7 (1%) | 9 (2%) |
| COVID-19 exposure | 29 (6%) | 40 (8%) |
| Quarantined | 212 (42%) | 88 (18%) |
| Practicing physical distancing | 501 (99%) | 490 (98%) |
| In voluntary self-isolation | 221 (44%) | 151 (30%) |
| In mandatory self-isolation | 54 (11%) | 33 (7%) |
| Working from home | 327 (65%) | 254 (51%) |
| Shelter-in-place/stay-at-home-order | 427 (84%) | - |
| Family member physically ill from COVID-19 | 35 (7%) | 67 (13%) |
| Family member hospitalized because of COVID-19 | 9 (2%) | 18 (4%) |
| Family member in isolation/quarantine | 131 (26%) | 71 (14%) |
| Family member lost job because of COVID-19 | 126 (25%) | 33 (6%) |

Percentages on available data are given when there are missing data. Values are *N*s and percentages.

#### COVID-19-related impact on eating disorder illness

We describe the results for the whole sample combining those who endorsed “somewhat” or “very concerned” on items about eating disorder concerns (Table 3) and “frequently” or “daily or more” on items about eating disorder illness behaviors (Table 4). The most prevalent concerns were similar in both countries. Foremost, 79% (US, *N* = 397) and 66% (NL, *N =* 331) of respondents were concerned about worsening of the eating disorder due to lack of structure. Furthermore, respondents were concerned about worsening of the eating disorder due to being in a triggering environment (US 58%; NL 57%) or lack of social support (US 59%; NL 48%), and being unable to access food consistent with their meal plan (US 61%; NL 36%). Concerns about having access to enough food or not being able to afford food or treatment were less commonly endorsed in both countries. Descriptive data showed that the greatest impact of COVID-19 on eating disorder behaviors for US participants was in the domain of feeling anxious about not being able to exercise (57%). This question was not asked of NL participants. Over one third of participants in both countries reported worsening of dietary restriction and compensatory behaviors. Regular binge eating on stockpiled food was reported by 23% of respondents in the US and 14% in NL.

**Table 3.**
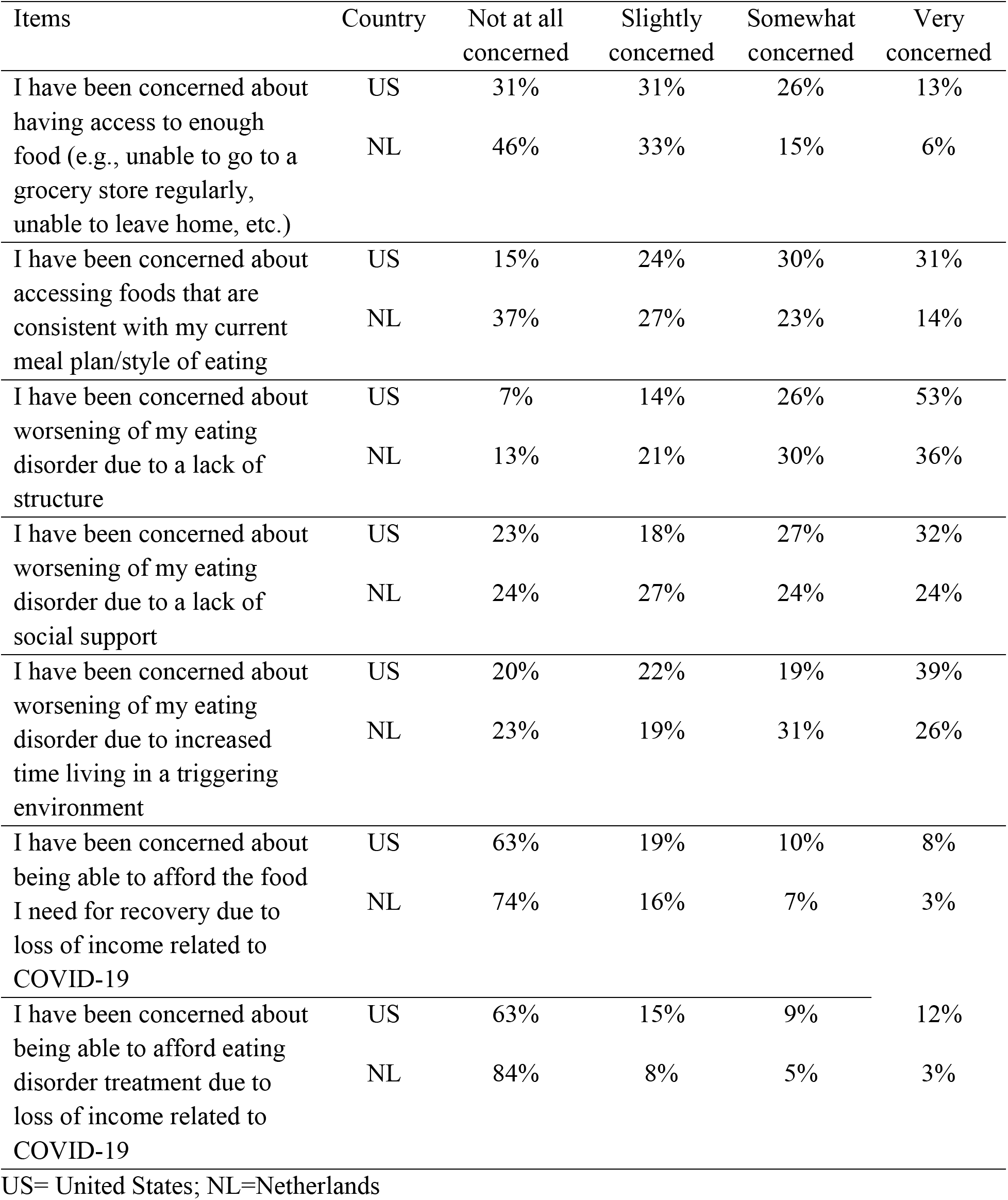
Concerns about impact of COVID-9 on eating disorder (United States: *N* = 511, Netherlands: *N* = 499)

| Items | Country | Not at all concerned | Slightly concerned | Somewhat concerned | Very concerned |
| --- | --- | --- | --- | --- | --- |
| I have been concerned about having access to enough food (e.g., unable to go to a grocery store regularly, unable to leave home, etc.) | US | 31% | 31% | 26% | 13% |
|  | NL | 46% | 33% | 15% | 6% |
| I have been concerned about accessing foods that are consistent with my current meal plan/style of eating | US | 15% | 24% | 30% | 31% |
|  | NL | 37% | 27% | 23% | 14% |
| I have been concerned about worsening of my eating disorder due to a lack of structure | US | 7% | 14% | 26% | 53% |
|  | NL | 13% | 21% | 30% | 36% |
| I have been concerned about worsening of my eating disorder due to a lack of social support | US | 23% | 18% | 27% | 32% |
|  | NL | 24% | 27% | 24% | 24% |
| I have been concerned about worsening of my eating disorder due to increased time living in a triggering environment | US | 20% | 22% | 19% | 39% |
|  | NL | 23% | 19% | 31% | 26% |
| I have been concerned about being able to afford the food I need for recovery due to loss of income related to COVID-19 | US | 63% | 19% | 10% | 8% |
|  | NL | 74% | 16% | 7% | 3% |
| I have been concerned about being able to afford eating disorder treatment due to loss of income related to COVID-19 | US | 63% | 15% | 9% | 12% |
|  | NL | 84% | 8% | 5% | 3% |

US= United States; NL=Netherlands

**Table 4.** Impact of COVID-19 on eating disorder behaviors (United States: *N* = 511, Netherlands: *N* = 492)

| Items | Country | Not<br>at<br>all | Once<br>or<br>twice | Frequently | Daily<br>or<br>more |
| --- | --- | --- | --- | --- | --- |
| In the past two weeks, I have binged on food that I (or my family or roommate) have stockpiled | US | 51% | 26% | 17% | 6% |
|  | NL | 71% | 15% | 9% | 5% |
| In the past two weeks, I have restricted my intake more because of COVID-19-related factors | US | 23% | 28% | 29% | 19% |
|  | NL | 36% | 24% | 25% | 14% |
| In the past two weeks, I have engaged in more compensatory behaviors (e.g., self-induced vomiting, excessive exercise, misuse of laxatives and/or water pills) because of COVID-19-related factors | US | 43% | 22% | 20% | 15% |
|  | NL | 38% | 24% | 23% | 15% |
| In the past two weeks, I have felt anxious about not being able to exercise | US | 18% | 25% | 29% | 28% |
|  | NL | - | - | - | - |

Netherlands participants were not asked the question about exercise.
US=United States; NL=Netherlands

Individuals with past eating disorders (i.e., not currently symptomatic) (US *N* = 37; NL *N* = 39) also expressed concerns. These participants reported concerns in a number of areas: worsening of the eating disorder due to a lack of structure (US 30%; NL 28%), increased time spent in a triggering environment (US 30%; NL 10%), access to food consistent with meal plan (US 32%; NL 3%), and having access to enough food (US 24%; NL 3%). They also reported worsening and frequent anxiety about exercise (US 35%; not asked in NL), restriction of intake (US 13%; NL 13%), and compensatory behaviors (US 5%; NL 3%).

##### Comparisons in lifetime diagnostic subgroups

In both countries, many group differences emerged on the eating disorder concern and illness behavior questions that were generally consistent with diagnostic characteristics (*P*s < 0.05). For example, the AN group reported significantly greater concerns about accessing foods consistent with their meal plan and a greater worsening of dietary restriction. The BED and BN groups reported more frequent binge eating of stockpiled food. The results on the individual items are shown in Supplementary Tables 1 and 2.

##### Comparisons by treatment status

In the US, participants who reported difficulty accessing their treatment provider reported higher mean eating disorder impacts (*N* = 32, *M* = 2.69, *SD* = 0.52) compared to those who reported that they had received face-to-face or online/tele-health treatment (*N* = 243, *M* = 2.40, *SD* = 0.58), *F*(1,273) = 6.94, *P* = 0.01. In NL, there were no differences between these groups (difficulty assessing treatment: *N* = 24, *M* = 2.08, *SD* = 0.62; still receiving care: *N* = 235, *M* = 2.13, *SD* = 0.53), *F*(1, 252) = 0.12, *P* = 0.91.

##### Other comparisons

Comparisons of eating disorder symptomatology by illness status (currently ill versus individuals with past eating disorders) and level of lockdown can be found in the Supplementary Material.

#### COVID-19-related impact on general physical and mental well-being

Table 5 presents items on the impact of COVID-19 on physical and mental health and any positive changes that have occurred. The majority of US and NL participants were somewhat to very worried about the impact of COVID-19 on their physical health (US 70%; NL 66%), but more reported concerns about the impact of COVID-19 on their mental health (US 86%; NL 88%). The mean score on the five COVID-19 worry items was 4.56 (*SD* = 1.24) for the US sample and 4.52 (*SD* = 1.17) in the NL sample which were between the scale anchors 4 = “somewhat worried” and 7 = “very worried”. Half of the US participants and ~40% of the NL participants reported that the COVID-19 situation had led to positive changes in their life.

**Table 5.** Impact of COVID-19 on general physical and mental well-being (United States: *N* = 511, Netherlands: *N* = 471)

| Items | Country | Response options |  |  |  |  |  |  |
| --- | --- | --- | --- | --- | --- | --- | --- | --- |
| How worried are you about being infected yourself? |  | 1 = Not worried at all | 2 | 3 | 4 = Somewhat worried | 5 | 6 | 7 = Very worried |
|  | US | 14% | 17% | 10% | 27% | 13% | 8% | 11% |
|  | NL | 20% | 17% | 8% | 21% | 20% | 10% | 5% |
| How worried are you about others being infected? | US | 2% | 3% | 5% | 24% | 16% | 20% | 30% |
|  | NL | 3% | 4% | 4% | 13% | 32% | 26% | 19% |
| How worried are you that your physical health could be influenced by coronavirus (COVID-19)? | US | 9% | 11% | 10% | 28% | 16% | 10% | 16% |
|  | NL | 13% | 13% | 7% | 19% | 19% | 16% | 12% |
| How worried are you that your mental health could be influenced by coronavirus (COVID-19)? | US | 3% | 5% | 6% | 13% | 15% | 21% | 37% |
|  | NL | 4% | 3% | 3% | 10% | 26% | 25% | 27% |
| How much of your day are you thinking about COVID-19? |  | 1 = None at all | 2 | 3 | 4 = About half the day | 5 | 6 | 7 = All day long |
|  | US | 1% | 17% | 22% | 23% | 16% | 14% | 6% |
|  | NL | 0.2% | 9% | 19% | 25% | 25% | 15% | 7% |
| Has the COVID-19 situation led to any positive changes in your life? |  | 1 = Not at all | 2 | 3 | 4 = Some positive changes | 5 | 6 | 7 = Several positive changes |
|  | US | 19% | 17% | 15% | 35% | 6% | 3% | 5% |
|  | NL | 28% | 16% | 17% | 18% | 16% | 4% | 1% |

The percentage of individuals screening positive for GAD was 68% (*N* = 328) in the US and 63% (*N* = 296) in NL. Mean GAD scores were 12.61 (*SD* = 5.60) and 11.83 (*SD* = 5.41), respectively. The majority of participants (US 80%; NL 65%) reported an increase in anxiety levels since the end of 2019, and only 0.2% (US) and 3% (NL) of this group reported that this change in anxiety was not at all due to COVID-19.

#### COVID-19-related impact on eating disorder treatment

Table 6 presents treatment status over the past 2 weeks (Table 6). Most respondents had transitioned to online/tele-health care (US 45%; NL 42%), with smaller numbers still receiving face-to-face care (US 3%; NL 6%), or not having been able to engage with their provider at all (US 6%; NL 5%). Consistent with the literature, high numbers of respondents were not receiving any eating disorders treatment (US 45%; NL 47%). Comparisons of the impact of COVID-19 on eating disorder treatment by diagnostic subgroup and level of lockdown can be found in the Supplementary Material.

**Table 6.** Impact of COVID-19 on eating disorder treatment (United States: *N* = 511, Netherlands: *N* = 488)

| Items | Country | Response options |  |  |  |
| --- | --- | --- | --- | --- | --- |
| Choose the best alternative that characterizes your situation over the past two weeks? |  | I have had face-to-face (in person) interactions with my eating disorders treatment provider(s) | I have transitioned to online care with my eating disorders treatment provider(s) (i.e., telehealth) | I have not been able to engage with my eating disorders treatment provider(s) at all | I do not currently receive eating disorders treatment |
|  | US | 3% | 45% | 6% | 45% |
|  | NL | 6% | 42% | 5% | 47% |
| The quality of my treatment in the past two weeks has been: |  | Better than usual | As good as usual | Somewhat worse than usual | Much worse than usual |
|  | US | 5% | 48% | 40% | 7% |
|  | NL | 4% | 22% | 56% | 18% |
| In the last two weeks: |  | I have had to reduce the number of sessions/contacts with my eating disorders treatment provider(s) |  | I have had at least the same number of sessions/contacts with my eating disorders treatment provider(s) |  |
|  | US | 23% |  | 77% |  |
|  | NL | 35% |  | 65% |  |

US= United States; NL= Netherlands

##### Comparisons in currently ill versus past ill individuals

In both countries, participants reporting a current eating disorder (US *N* = 474, 93%; NL *N* = 468, 92%) had a significantly higher mean worry score (US *F*(1, 484) = 5.28, *P* = 0.02); NL (*F*(1, 464) = 5.96, *P* = 0.02) than those with past eating disorders. The mean worry scores were 4.59 (*SD* = 1.24) for those with a current eating disorder versus 4.11 (*SD* = 1.22) (US) for those with a past eating disorder—in NL, these scores were 4.56 (*SD* = 1.17) versus 4.05 (*SD* = 1.01). Likewise, those with a current eating disorder had a significantly higher GAD-7 score (US *F*(1, 481) = 11.78, *P* < 0.001; NL *F*(1, 460) = 16.79, *P* < 0.001) with a mean of 12.87 (*SD* = 5.54) versus 9.62 (*SD* = 5.36) (NL scores were 12.10 [*SD* = 5.37] versus 8.43 [*SD =* 4.81]). The percentages screening positive for GAD were 70% (US) and 66% (NL) of participants with current eating disorders compared with 41% (US) and 37% (NL) of those with past eating disorders (US χ^2^(1) = 13.43, < 0.001; NL χ^2^ (1) = 11.23, < 0.001).

##### Comparisons by treatment status

Among those who were receiving treatment, there were no significant differences in mean worry score between those who reported that they had not been able to engage with their treatment provider at all (US *N* = 29, 11%; NL *N* = 24, 9%) versus those who reported continuity of care in the form of face-to-face or online/tele-health treatment (US *N* = 239, 89%; NL *N* = 235, 91%), (US *F*(1,266) = 0.01, *P* = 0.93; NL *F(*1,244) = 0.18, *P* = 0.67). The mean worry score was 4.63 (*SD* = 1.18) for those having difficulty with treatment access versus 4.65 (*SD* = 1.19) for those reporting continuity of treatment, and 4.7 (*SD* = 1.14) versus 4.5 (*SD* = 1.19) in NL. There were no significant differences in GAD-7 scores in both countries (US *F*(1,264) = 0.01, *P* = 0.93); NL *F*(1, 241) = 1.49, *P* = 0.22). The mean GAD-7 score was 12.86 (*SD* = 6.19) for those having difficulty with treatment access versus 12.96 (*SD* = 5.41) for those reporting continuity of care, and 11.14 (*SD* = 5.67) versus 12.66 (*SD* = 5.30) in NL.

Comparisons of the impact of COVID-19 on general physical and mental well-being by lifetime diagnostic subgroups and level of lockdown can be found in Supplementary Material.

### QUALITATIVE RESULTS

A series of open-ended survey questions asked participants to describe a) any positive changes in their eating disorder; b) treatment needs; c) other concerns not queried elsewhere in the survey; and d) the any positive changes the COVID-19 situation had produced in their lives. Three authors (CB, CP, EvF) reviewed the qualitative responses and identified broad themes within the responses for each question (see Table 7 and 8). Given our goal of providing strictly descriptive results, we did not undertake formal efforts at establishing a coding scheme.

**Table 7.**
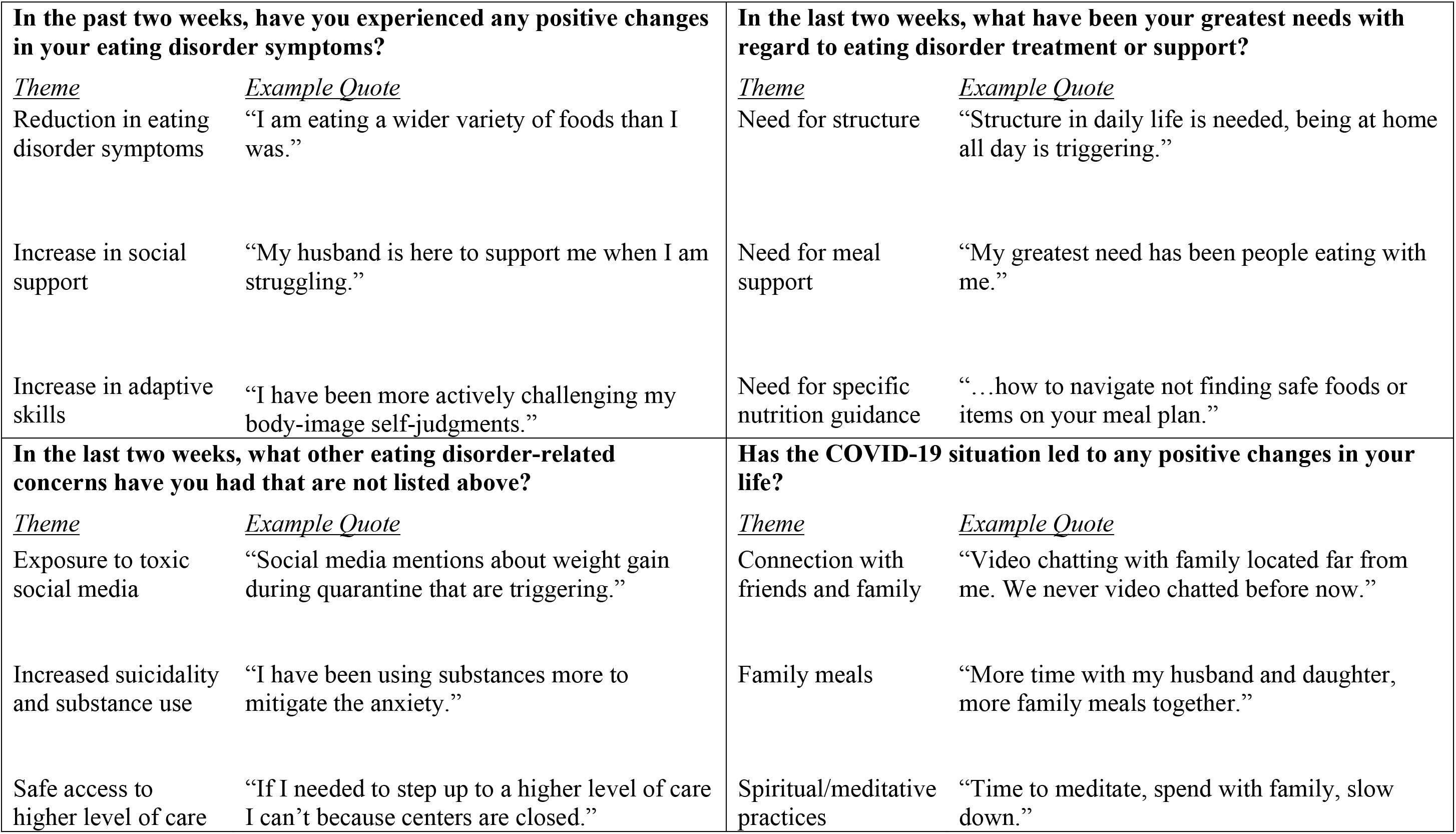
Summary of themes obtained from qualitative US data.

**Table 8.**
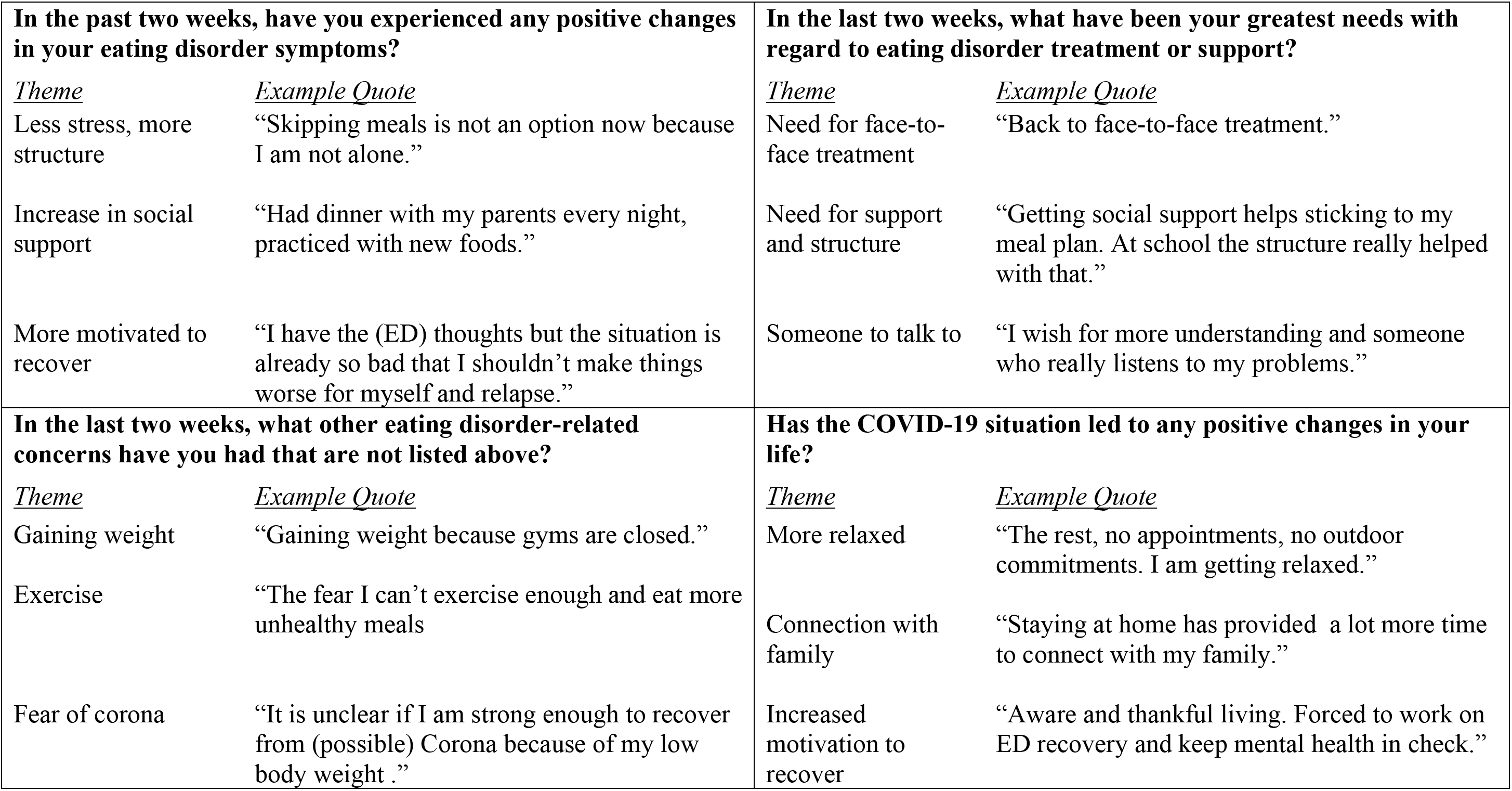
Summary of themes obtained from qualitative Netherlands data.

A positive consequence of the changes due to COVID-19 reported by respondents was a perceived increase in social support that helped challenge their eating disorder behaviors and increase motivation to recover. Their greatest treatment need was for more structure, face-to-face treatment and someone to talk to (NL only), meal support and nutritional guidance (US only). Other eating disorder-related concerns differed by country. In the US, exposure to toxic social media, increased suicidality and substance use, and safe access to higher levels of care were dominant themes. In NL, respondents’ main concerns were fear of gaining weight, increased desire to exercise, and a fear of what would happen were they to contract COVID-19. In terms of positive changes in their life, participants in both countries valued the increased connection with family and friends. The Dutch participants highlighted feeling more relaxed and an increased motivation to work on recovery. Family meals and spiritual and meditative practices were a main theme mentioned by US respondents.

## DISCUSSION

We describe the early impact of COVID-19 and treatment needs in patients with eating disorders in the US and NL. At the time of the study launch, the US had been in some degree of lockdown for approximately three to five weeks. In the NL restrictions had been underway for approximately one month. Although fairly few participants were directly affected by COVID-19, the overwhelming majority were indirectly affected, anxiety levels were elevated, and concerns were high regarding the impact of COVID-19-related factors on their eating disorder and on their mental health in general.

Consequences of the lockdown measures—a lack of structure, increased time spent in a triggering environment, lack of social support—were common concerns. Additionally, worsening of eating disorder behaviors was broadly consistent with respondents’ self-reported eating disorder diagnosis. US participants were particularly anxious about not being able to exercise. Of note, even those who were not currently symptomatic endorsed some degree of concern indicating a heightened vulnerability for relapse.

Our results align with other studies showing worsening anxiety during COVID-19.^1^ Mean GAD-7 scores were substantially higher than typical in the general population during non-pandemic circumstances^20^ (*N* = 5030, *M* = 2.97, 95 CI = 2.86-3.07), but on par with scores in both general psychiatric disorder and eating disorder samples.^21,25^ To our knowledge, GAD-7 has not been assessed in the general population in the US or NL during the COVID-19 pandemic for comparison.

Encouragingly, more than one-third (US 49%; NL 40%) of participants identified that COVID-19 had led to positive changes in their life. Many reported a sense of connection with family and friends, an ability to focus on recovery-oriented goals, and engagement in adaptive coping skills. That respondents were able to name positive effects of this pandemic in addition to acknowledging the deleterious effects it has had on their eating disorder highlights the complex nature of individuals’ experiences.

We were particularly interested in ascertaining participants’ perceptions of treatment during COVID-19. Unfortunately, nearly half of both samples reported not currently receiving treatment for their eating disorder (despite the majority endorsing current symptoms). This is consistent with many previous reports indicating that eating disorders are often underdetected and undertreated.^26^ For those who were in care, most had transitioned to telehealth services at a similar frequency to previous face-to-face sessions. However, the transition was not without challenges. In our sample, 47% of US respondents and 74% of NL respondents reported that the quality of their treatment had been “somewhat” or “much” worse than usual. It is not clear what influenced this perception, and we note that the survey was deployed at a time when many clinicians were just making the transition to remote care. Follow-up surveys will allow us to address whether quality of remote care continues to be a concern.

To our knowledge, this is the first large-scale study to capture concerns of individuals with eating disorders during COVID-19. Our rapid deployment did yield limitations. First, eating disorder diagnoses were self-reported as interviews were not feasible in the timeframe. Second, despite many striking similarities, differences between the US and NL emerged in terms of sample composition as well as the countries’ approach to pandemic control—with the NL strategy being more comprehensive and uniform and the US being more fragmented. Moreover, differences in these strategies and stage of the pandemic at deployment could affect the pattern of responses. Our use of convenience sampling could introduce bias in the responses, as individuals who were most worried/concerned might be more inclined to participate. Finally, our use of predetermined questions, did not allow for an unbiased survey of respondents’ concerns. Although outside of the scope of the present study, given that many individuals with eating disorders returned to their families of origin during lockdown, it would be of value to document the impact of COVID-19 on carer’s and family member’s experiences particularly with relation to stress, burnout, mealtimes, and healthcare provider support.

In summary, individuals with eating disorders may be experiencing a worsening of symptoms and that those with past eating disorders may be vulnerable for relapse during COVID-19. Although these data are primarily descriptive, we aim to provide preliminary guidance to healthcare providers about ways in which they might be of assistance during this time. Both quantitative and qualitative data underscore the call for structure, especially around meals. Clinicians should assess what structures do exist and be innovative in developing strategies to help patients structure their daily schedules and meals.

Given the unclear level of satisfaction with telehealth approaches, clinicians should be direct in addressing what is working and what is not working in terms of telehealth sessions, and not assume that they are equivalent to face to face encounters. Frank discussions might allow patient and provider to work collaboratively to ensure care is being delivered effectively and compassionately. Finally, given the uncertain nature of this pandemic and the ways in which it will affect our lives, it is important to remain nimble both in the approach to delivering care and the extent to which care may need to be modified in light of novel and ever-changing stressors.

Assessing and harnessing factors that have led to positive changes in patients’ lives and their eating disorder, such as greater social support, family mealtimes, and opportunities for self-care might may aid with treatment planning. The challenges in delivering eating disorders care during COVID-19 are numerous; however, consistent collaborative efforts between patient and provider may help stem the tide of worsening symptomatology and allow patients to continue to make progress toward recovery.

## Data Availability

Anonymized data will be available at the end of the final follow-up survey (12 months after the baseline study) by contacting the corresponding author.

## Acknowledgements

Dr Bulik acknowledges support from the National Institutes of Health (R01MH120170; R01MH119084) and the Swedish Research Council (Vetenskapsrådet, award: 538-2013-8864). Drs Peat and Bulik acknowledge support from the Substance Abuse and Mental Health Administration (H79 SM081924).

## Conflicts

CM Bulik reports: Shire (grant recipient, Scientific Advisory Board member); Idorsia (consultant); Pearson (author, royalty recipient); CM Peat reports: Sunovion (Scientific Advisory Board member).

## Notes

### Funding Statement

The research presented in the paper did not receive external funding.
The authors report the following general support: Dr Bulik acknowledges support from the National Institutes of Health (R01MH120170; R01MH119084) and the Swedish Research Council (538-2013-8864). Drs Peat and Bulik acknowledge support from the Substance Abuse and Mental Health Administration (H79 SM081924).

### Author Declarations

Ethical approval was granted by the University of North Carolina Biomedical Institutional Review Board. The Medical Research Ethics Committee (MREC-LDD) of Leiden University Medical Centre reviewed the study protocol and confirmed that the Medical Research Involving Human Subjects Act (WMO) did not apply to this study and official approval of this study by the METC was not required.

## References

1. Fowers A, Wan W. A third of Americans now show signs of clinical anxiety or depression, Census Bureau finds amid coronavirus pandemic. Washington Post. 2020 May 26 2020.

2. Adhanom Ghebreyesus T. Addressing mental health needs: an integral part of COVID-19 response. World Psychiatry. 2020;19:129–30.

3. Twenge J, Joiner TE. Mental distress among U.S. adults during the COVID-19 pandemic. PSsyArXiv. 2020.

4. The Lancet Psychiatry. Mental health and COVID-19: change the conversation. Lancet Psychiatry. 2020.

5. Kaufman KR, Petkova E, Bhui KS, Schulze TG. A global needs assessment in times of a global crisis: world psychiatry response to the COVID-19 pandemic. BJPsych Open. 2020:1-11.

6. Galletly C. Psychiatry in the COVID-19 Era. Aust N Z J Psychiatry. 2020;54(5):447–8.

7. Hao F, Tan W, Jiang L, Zhang L, Zhao X, Zou Y, et al. Do psychiatric patients experience more psychiatric symptoms during COVID-19 pandemic and lockdown? A case-control study with service and research implications for immunopsychiatry. Brain Behav Immun. 2020.

8. Liu N, Zhang F, Wei C, Jia Y, Shang Z, Sun L, et al. Prevalence and predictors of PTSS during COVID-19 outbreak in China hardest-hit areas: Gender differences matter. Psychiatry Res. 2020;287:112921.

9. Wang C, Pan R, Wan X, Tan Y, Xu L, McIntyre RS, et al. A longitudinal study on the mental health of general population during the COVID-19 epidemic in China. Brain Behav Immun. 2020.

10. Zhou J, Liu L, Xue P, Yang X, Tang X. Mental health response to the COVID-19 outbreak in China. Am J Psychiatry. 2020:appiajp202020030304.

11. Fernández-Aranda F, Casas M, Claes L, Bryan DC, Favaro A, Granero R, et al. COVID-19 and implications for eating disorders. Eur Eat Disord Rev. 2020;28:239.

12. Touyz S, Lacey H, Hay P. Eating disorders in the time of COVID-19. J Eat Disorders. 2020;8:19.

13. Weissman RS, Bauer S, Thomas JJ. Access to evidence-based care for eating disorders during the COVID-19 crisis. Int J Eat Disord. 2020.

14. Davis C, Chong NK, Oh JY, Baeg A, Rajasegaran K, Chew CSE. Caring for children and adolescents with eating disorders in the current COVID-19 pandemic: A Singapore perspective. J Adol Health. 2020.

15. Murphy R, Calugi S, Cooper Z, Dalle Grave R. Challenges and opportunities for enhanced cognitive behaviour therapy (CBT-E) in light of COVID-19. Cogn Behav Ther. 2020:1-31.

16. Waller G, Pugh M, Mulkens S, Moore E, Mountford VA, Carter J, et al. Cognitive-behavioral therapy in the time of coronavirus: Clinician tips for working with eating disorders via telehealth when face-to-face meetings are not possible. Int J Eat Disord. 2020.

17. Thornton LM, Munn-Chernoff MA, Baker JH, Jureus A, Parker R, Henders AK, et al. The Anorexia Nervosa Genetics Initiative (ANGI): Overview and methods. Contemp Clin Trials. 2018;74:61–9.

18. Bulik CM, Butner J, Tregarthen J, Thornton LM, Flatt R, Smith T, et al. The Binge Eating Genetics Initiative (BEGIN) Study. BMC Psychiatry. 2020.

19. Spitzer RL, Kroenke K, Williams JB, Lowe B. A brief measure for assessing generalized anxiety disorder: the GAD-7. Arch Intern Med. 2006;166:1092–7.

20. Löwe B, Decker O, Müller S, Brähler E, Schellberg D, Herzog W, et al. Validation and standardization of the Generalized Anxiety Disorder Screener (GAD-7) in the general population. Med Care. 2008;46:266–74.

21. Weigel A, Konig HH, Gumz A, Lowe B, Brettschneider C. Correlates of health related quality of life in anorexia nervosa. Int J Eat Disord. 2016;49:630–4.

22. Strandskov SW, Ghaderi A, Andersson H, Parmskog N, Hjort E, Warn AS, et al. Effects of tailored and ACT-influenced internet-based CBT for eating disorders and the relation between knowledge acquisition and outcome: A randomized controlled trial. Behav Ther. 2017;48:624–37.

23. Weigel A, Lowe B, Kohlmann S. Severity of somatic symptoms in outpatients with anorexia and bulimia nervosa. Eur Eat Disord Rev. 2019;27:195–204.

24. Schaumberg K, Jangmo A, Thornton L, Birgegård A, Almqvist C, Norring C, et al. Patterns of diagnostic flux in eating disorders: a longitudinal population study in Sweden. Psychol Med. 2019;49:432–50.

25. Donker T, van Straten A, Marks I, Cuijpers P. Quick and easy self-rating of Generalized Anxiety Disorder: validity of the Dutch web-based GAD-7, GAD-2 and GAD-SI. Psychiatry Res. 2011;188(1):58–64.

26. Hart LM, Granillo MT, Jorm AF, Paxton SJ. Unmet need for treatment in the eating disorders: a systematic review of eating disorder specific treatment seeking among community cases. Clin Psychol Rev. 2011;31(5):727–35.

